# Rx-LLM: a benchmarking suite to evaluate safe large language model performance for medication-related tasks

**DOI:** 10.64898/2025.12.01.25341004

**Authors:** Xingmeng Zhao, Kaitlin Blotske, Moriah Cargile, Adeleine Tilley, Brian Murray, Yanjun Gao, Kelli Henry, Susan E. Smith, Erin F. Barreto, Seth Bauer, Sunghwan Sohn, Tianming Liu, Tell Bennett, Mitch Cohen, Andrea Sikora

## Abstract

**Background:** For large language models (LLMs) to reach their potential as information technology tools that make medication use safer, clinically relevant benchmarks capable of automated grading and designed specifically to measure the performance of LLMs for medication tasks are required. The purpose of this study was to design a suite of benchmarking tests reflective of Comprehensive Medication Management (CMM; the standard of care for medication optimization) and quantify the baseline performance of the latest LLMs.

**Methods:** We established six benchmarks representing critical stages of the CMM process: drug formulation matching, drug order (sig) generation, drug route matching, drug-drug interaction identification, renal dose identification, and drug-indication matching. For each benchmark, we curated a clinician-annotated dataset comprising 250 standardized input-output pairs including both inpatient and outpatient medications. We evaluated the clinical knowledge retrieval capabilities of three LLMs: GPT-4o-mini, MedGemma-27B, and LLaMA3-70B. We employed a zero-shot prompting strategy, excluding in-context examples, to assess the models’ internal clinical knowledge rather than their few-shot learning potential. To check reliability, each model was run three times using a temperature of 0.7 (a mid-range value of an LLM setting controlling text generation randomness). Performance was assessed using task-specific evaluation metrics including precision (positive predictive value), recall (sensitivity), F1-score, accuracy, and correctness consistency across trials.

**Results:** Across six benchmarks, **LLaMA3-70B** demonstrated the highest performance in four tasks: drug-formulation matching (F1, 54.0% [95 CI: 50.1-58]), drug-order generation (accuracy, 88.0%), drug-route identification (F1, 74.3% [95 CI: 71-78]), and drug-indication identification (accuracy, 97.6% [95 CI: 95.6-99.2]). In the drug–drug interaction task, **GPT-4o-mini** achieved the highest overall accuracy (70.4% [95 CI: 64.8-75.7]). For renal dose–adjustment identification, GPT-4o-mini demonstrated the highest F1 score (83.3% [95 CI: 77.6-88]). Correctness-consistency scores ranged from 8.0% to 97.6% across benchmarks, with no model exhibiting uniformly superior consistency.

**Conclusions:** Model performance varied substantially across medication-related tasks. LLaMA3-70B demonstrated promising baseline performance in tasks involving formulation, ordering, route, and indication. GPT-4o-mini showed potential advantages in drug–drug interaction detection and renal dose adjustment. These findings underscore the need for task-specific evaluation when deploying models for medication-focused clinical decision support.

## Background

Large language models (LLMs) have demonstrated potential for use as information technology (IT) tools in the space of comprehensive medication management (CMM). However, the use of LLMs for medication-related tasks requires standardized benchmarks that have the capability of being automated to avoid resource intensive clinician annotation to ensure safety and efficacy.^1–4^

Medications represent both a unique knowledge domain for LLMs and a construct far removed from diagnosis-related tasks. In disease diagnosis, the ground truth is readily available in large, open-source datasets (e.g., radiology images corresponding to a patient’s cancer diagnosis). Treatment is a far less structured problem wherein possible therapies have a range of potential characteristics (e.g., highly safe and efficacious, reasonably efficacious, safe but inefficacious, efficacious but harmful, harmful and inefficacious). Most treatment decisions have multiple “right” answers, a range of acceptable answers, and “wrong” answers that range from benign to deadly. The complexities of medication treatment are managed by interprofessional and interdisciplinary teams including clinical pharmacists who are charged with performing CMM. CMM is defined as “the standard of care that ensures each patient’s medications are individually assessed to determine that each medication is appropriate for the patient, effective for the medical condition, safe given the comorbidities and other medications being taken, and able to be taken by the patient as intended.”^5^

Similar to drug therapy, which undergoes Food and Drug Administration (FDA) approval for safety and efficacy outcomes, LLMs require careful evaluation for their ability to provide information that is safe from harm and has the potential for benefit. However, an FDA perspective on AI regulation states “The sheer volume of these changes and their impact also suggests the need for industry and other external stakeholders to ramp up assessment and quality management of AI across the larger ecosystem beyond the remit of the FDA” and underscores that “all involved sectors will need to attend to AI with the care and rigor this potentially transformative technology merits”.^6,7^ This is particularly important given that LLMs have shown vulnerability to poisoning and hallucinations, which could result in life-threatening medication regimen recommendations.^8,9^

To date, no organization or institution has undertaken the necessary task of establishing benchmarks for the safe performance of LLMs for clinical acceptance in the domain of medication tasks. The purpose of this study was to design a suite of benchmarks reflective of the CMM process to allow for the performance evaluation of LLM-based IT tools.

## Methods

### Study design

A panel of 5 licensed, board-certified critical care pharmacists with a range of practice settings (academic medical centers, community hospitals, etc.) was convened to review the best practices for CMM in the ICU based on published consensus recommendations and guidelines.^10–12^ The panel developed a visual diagram of the reasoning process for clinical decision-making within the CMM process (see **Figure 1**). Tasks were selected to capture a range of clinical reasoning skills, medication knowledge, and structured decision outputs relevant to real-world prescribing. Each task was formulated to require direct model generation or classification without access to external tools.

**Figure 1.**
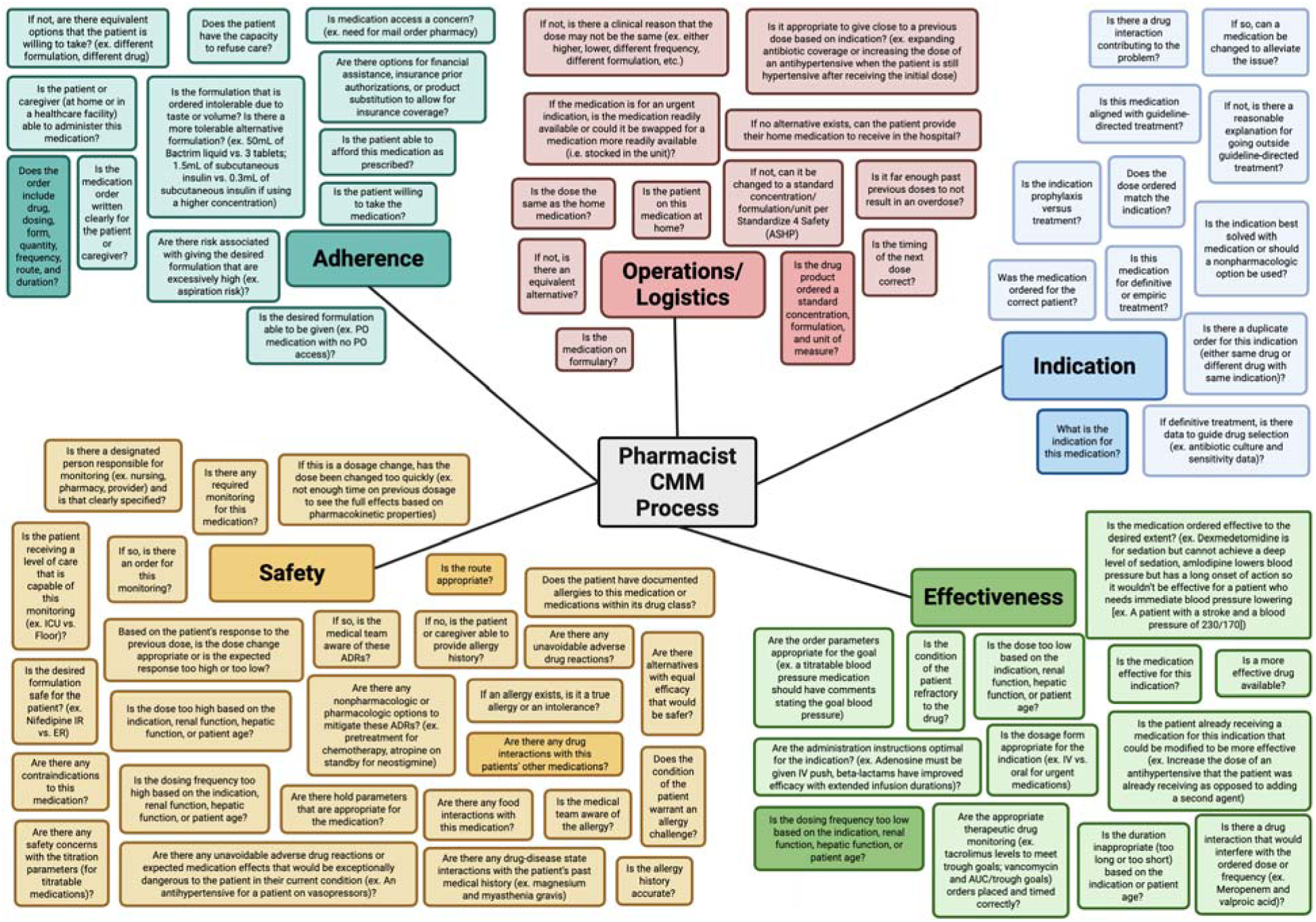
Comprehensive Medication Management Reasoning Process Comprehensive medication management (CMM) is a complex reasoning process including domain specific knowledge that integrates knowledge of pharmacotherapy, pharmaceutics, institution specific standards, disease state management, and patient specific information. This figure represents important questions asked and answered throughout that process. Darker shading represents questions that are specifically evaluated in RxLLM.

We conducted a structured evaluation of three LLMs. LLaMA3-70B, GPT-4o-mini, and MedGemma-27B, across six medication-related benchmarks designed to reflect common and safety-critical tasks in clinical medication management. These LLMs were selected due to their widespread availability. The panel identified six benchmark tasks at key junctures in the CMM process representing various medication-related tasks from the major categories of CMM (safety, adherence, operations, indication, effectiveness). The primary input concept and output concept are mapped in **Table 1**.

**Table 1.** Input and output pairs for the six comprehensive medication management benchmark tasks (all n=250)

| Benchmark Task | Description | Input | Output |
| --- | --- | --- | --- |
| **Drug formulation matching** | Tests the ability to correctly match the generic drug name with the available dosage forms | Generic drug name (e.g., amlodipine) | All FDA-approved formulations available (e.g., 2.5 mg tablet, 5 mg tablet, 10 mg tablet, 1 mg/mL oral solution, 1 mg/mL oral suspension) |
| **Drug order (sig) generation** | Tests the ability to generate a safe and appropriate complete medication order given a drug name | Generic drug name (e.g., carvedilol) | One complete order for an oral route (e.g., carvedilol 12.5 mg, one tablet by mouth twice daily) |
| **Drug route matching** | Tests the ability to identify all safe routes (e.g. intravenous, oral, ophthalmic) available for a medication. | Generic drug name (e.g., prednisolone) | All routes available (e.g., oral, ophthalmic) |
| **Drug-drug interaction identification** | Tests the ability to correctly identify which drugs have a drug-drug interaction given a list of 4-6 concomitant medications | List of 4-6 drugs with complete orders (e.g., Tenecteplase 22.5 mg administered as IV bolus; Labetalol 20 mg IV given over 1 minute; Aspirin 325 mg by mouth once daily; Atorvastatin 40 mg by mouth once daily; Lorazepam 0.5 mg every 4 to 6 hours as needed for anxiety) | Two drugs with drug-drug interaction Category C, D, or X* among the list (e.g., tenecteplase and aspirin) |
| **Renal dose identification** | Tests the ability to identify if the intravenous or oral formulation of the medication requires a renal dose adjustment or is contraindicated in renal failure | Generic drug name (e.g., vancomycin) | Renal dose adjustment required or contraindicated at certain renal function for any available routes (yes/no) |
| **Drug-indication matching** | Tests the ability to identify which medication, among a list of three medications, is being used for a specific indication | Generic drug name of 3 medications (e.g., azithromycin, docusate, norepinephrine) and an indication (e.g. community acquired pneumonia) | Medication in the given regimen that is being used for the specific indication (e.g. azithromycin) |

CMM: comprehensive medication management, FDA: Food and Drug Administration, IV: intravenous, kg: kilogram, mg: milligram, mL: milliliter
\*Drug interaction categories are defined as follows:
Category X — Avoid combination: This drug pair should generally not be used together because the clinical risk outweighs any potential benefit.
Category D — Consider therapy modification: The interaction is clinically relevant, and modification of therapy (dose adjustment, substitution, or precautions) should be considered.
Category C — Monitor therapy: An interaction is present. The combination may be used but requires monitoring by a healthcare professional.

This project was reviewed and approved by the University of Colorado Institutional Review Board (COMIRB #25-1631). All methods were performed in accordance with the ethical standards of the Helsinki Declaration of 1975. This evaluation followed the transparent reporting of a multivariable model for individual prognosis or diagnosis (TRIPOD–LLM) extension reporting frameworks, as applicable (Supplemental Appendices 1).^13^

### Benchmark descriptions

The benchmark tasks were designed to capture core elements of medication knowledge and prescribing practice. For drug formulation and route identification, models were asked to list all clinically valid formulations and routes of administration for each medication; both tasks required multilabel predictions because many medications have multiple acceptable forms and routes. The drug-order generation task evaluated a model’s ability to produce a complete prescription sig, including dose, route, and frequency, based on a medication prompt and clinical context, with outputs compared against an expert-validated reference.

The remaining tasks focused on clinical appropriateness and safety. For drug-indication identification, models judged whether a medication was indicated for a specific condition. Drug–drug interaction classification required identifying two medications that have a significant drug interaction out of a list of 4-6 medications. Renal dose–adjustment identification assessed whether a medication required dosage modification in the presence of renal impairment.

While not comprehensive for all medication tasks, these tasks were considered essential for safe performance of an LLM. Notably, the six tasks are already safely performed or checked by non-artificial intelligence IT tools (which do not run the risk of hallucinations and have transparent data sources and coding), such that AI would ultimately need to surpass performance in these areas to supersede existing safe standards of care.

### Data source

A total of six datasets were curated to test the proposed benchmark tasks with 250 input-output pairs per task for a total of 1500 pairs. These datasets included both inpatient and outpatient medications. These datasets are posted on Github: https://github.com/sikora07/AIChemist. Source material was taken from LexiDrug for each medication, with review by 2 board-certified pharmacists to ensure accuracy and clinical relevance.^14^

### Model Evaluation Framework

Our primary objective was to assess each model’s ability to reproduce the ground-truth clinical annotation for each task. Because the tasks differ in structure, we defined a primary evaluation metric per task based on the type of prediction required. For tasks where the model selects the correct answer from a small, instance-specific set of candidate options, such as drug-order, drug-indication, and drug–drug interaction identification, we used accuracy as the primary metric. These tasks require an exact-match selection among the provided drugs or drug pairs, and partial-credit metrics (e.g., precision/recall) are not meaningful in this single-choice setting. Accuracy therefore best reflects whether the model retrieved the correct clinical entity without hallucination. For tasks involving multi-label or asymmetric error structures, such as drug-formulation, drug-route, and renal dose-adjustment, we used F1-score as the primary metric, as it balances precision and recall when false positives and false negatives carry different clinical implications. Specifically, for formulation and route (multi-label), we computed set-based precision and recall; for renal dose adjustment (binary), we calculated precision, recall, F1-score, and accuracy from the confusion matrix. For drug order, we calculated accuracy based on a manual review of errors by two pharmacists. Additionally, we evaluated correctness consistency, which measures the fraction of runs where predictions were correct, averaged across all instances. We also reported 95% confidence interval across runs to quantify variability in model performance.

We evaluated three LLMs representing different architectural approaches and training strategies: GPT-4o-mini (OpenAI), a general-purpose model optimized for efficiency; MedGemma-27B (Google), a domain-specific model trained on biomedical literature; and LLaMA3-70B (Meta), a large-scale general-purpose model. Model selection was designed to assess whether domain-specific training or model scale provides advantages for medication management tasks.

All models were evaluated using zero-shot prompting without in-context examples to assess inherent clinical knowledge embedded during pretraining, rather than few-shot learning capabilities. This approach isolates model knowledge from prompt engineering effects and reflects real-world deployment scenarios where examples may be unavailable. Task-specific prompts were developed through iterative refinement with clinical pharmacists to ensure clarity, minimize ambiguity, and maintain clinical authenticity while avoiding leading language that could bias responses.

To quantify model reliability and prediction consistency—critical metrics for clinical deployment—each model was evaluated three times per instance using different random seeds. This repeated evaluation approach enables assessment of both accuracy (correctness of predictions) and consistency (reproducibility across runs), which directly impact patient safety in clinical settings.^15,16^ Prediction variability was quantified using standard deviation across runs, providing insight into model stability. This methodology aligns with uncertainty quantification frameworks for LLMs and reflects best practices for evaluating AI trustworthiness in healthcare applications.^17^

### Evaluation metrics

We evaluated model performance using commonly used metrics: precision, recall, F1 score, and accuracy. Additionally, the Harm Associated with Medication Error Classification (HAMEC) score was used to classify drug errors noted in the order generation benchmark and was validated by two clinical pharmacists.^18^ Given the hypothesis generating nature of this exploration, no attempt was made to calculate sample size.

### Classification metrics

For tasks with single-label outputs, evaluation was based on the four parts of the confusion matrix: true positives (TP), false positives (FP), false negatives (FN), and true negatives (TN). Precision measured how often the model’s positive predictions were correct (TP / [TP + FP]). Recall measured how often the model correctly identified the true positive cases (TP / [TP + FN]). The F1 score combined precision and recall into a single number that balances both. Accuracy measured how often the model was correct overall and was calculated as:

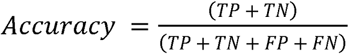

### Multilabel metrics

The drug-route and drug-formulation benchmark tasks required multilabel predictions, where a single medication could have more than one correct answer. For these tasks, we used two additional measures. Exact-match accuracy counted how often the model predicted every correct label for an item with no mistakes. Micro-averaged label accuracy looked across all labels and measured the proportion of labels the model predicted correctly out of all labels that appeared in either the predictions or the reference standard.

## Results

Across the six medication-related benchmark tasks, model performance differed substantially. As shown in **Table 2**, LLaMA3-70B achieved the highest performance for four tasks. In drug formulation matching, LLaMA3-70B produced the strongest F1 score (54.0% [95% CI: 50.1-58.0]), exceeding GPT-4o-mini (53.1% [95 CI: 49.0-57.0]) and MedGemma-27B (40.9% [95% CI: 36.5-45.2]). In drug order (sig) generation, LLaMA3-70B achieved the highest overall accuracy (88.0%) with fewer incorrect dose, incorrect frequency, and incomplete instruction errors than the other models. LLaMA3-70B also outperformed the other models in drug route identification (F1 74.3% [95% CI: 71.0-78.0]) and drug indication identification (97.6% [95% CI: 95.6-99.2]), as reported in **Tables 2 and 3**.

**Table 2.** Benchmarks 1-3: Drug Formulation Matching, Drug Order (sig) Generation, Drug Route Identification.

|  | **GPT-4o-mini (n=250)** | **MedGemma-27B (n=250)** | **LLaMA3-70B (n=250)** |
| --- | --- | --- | --- |
| ***Drug Formulation Matching*** |  |  |  |
| F1 | 53.1% [49.0, 57.0] | 40.9% [36.5, 45.2] | 54.0% [50.1, 58.0] |
| Recall | 43.0% [38.6, 47.4] | 33.0% [27.7, 38.2] | 49.5% [44.5, 54.2] |
| Precision | 69.6% [66.1, 73.0] | 54.0% [50.7, 57.3] | 59.5% [55.8, 63.1] |
| Exact Match Accuracy | 19.1% [14.7, 23.8] | 9.9% [6.4, 13.5] | 16.5% [12.1, 21.2] |
| Label Match Accuracy | 36.2% [32.5, 39.9] | 25.7% [22.3, 29.2] | 37.0% [33.5, 40.8] |
| Correctness Consistency Score, n (%) | 39 (15.6) | 20 (8.0) | 41 (16.4) |
| ***Drug Order (sig) Generation*** |  |  |  |
| Overall accuracy, n (%) | 208 (83.2) | 203 (81.2) | 220 (88.0) |
| Type of Error, n |  |  |  |
| Formulation Does Not Exist | 16 | 17 | 8 |
| Incorrect Dose | 9 | 12 | 10 |
| Incorrect Frequency | 12 | 16 | 10 |
| Incomplete Instructions | 15 | 14 | 6 |
| Harm Associated with Medication Error Classification (HAMEC Score), n (%) |  |  |  |
| 0 | 6 (2.4) | 3 (1.2) | 4 (1.6) |
| 1 | 19 (7.6) | 25 (10.0) | 10 (4.0) |
| 2 | 14 (5.6) | 17 (6.8) | 14 (5.6) |
| 3 | 2 (0.8) | 2 (0.8) | 3 (1.2) |
| 4 | 0 (0) | 1 (0.4) | 0 (0) |
| Did Not Follow Prompt Instructions, n (%) | 4 (1.6) | 17 (6.8) | 2 (0.8) |
| ***Drug Route Identification*** |  |  |  |
| F1 | 67.7% [64.4, 70.9] | 69.1% [65.6, 72.9] | 74.3% [71.0, 78.0] |
| Recall | 77.4% [73.1, 82.0] | 75.9% [71.6, 80.5] | 76.8% [72.7, 81.3] |
| Precision | 60.2% [56.4, 63.8] | 63.4% [59.2, 67.7] | 72.0% [67.5, 76.4] |
| Exact Match Accuracy | 39.7% [33.9, 45.2] | 49.7% [43.6, 55.9] | 56.7% [50.8, 63.1] |
| Label Match Accuracy | 51.2% [47.5, 55.0] | 52.8% [48.8, 57.3] | 59.1% [55.1, 63.9] |
| Correctness Consistency Score, n (%) | 81 (32.4) | 120 (48.0) | 139 (55.6) |

Values reported as mean [95% confidence interval] % of performance. Higher numbers indicate better performance.
Type of error reported as n, and some drugs may have multiple error types reported.
Recall: true positive/ (true positive + false negative). Low recall indicates many false negatives.
Precision: true positive/ (true positive + false positive). Low precision indicates many false positives
F1: 2 x (precision x recall)/(precision + recall). Low F1 indicates many false negatives, false positives, or both.
Overall accuracy defined as no errors present and followed prompt instructions for all 3 runs for the medication for each large language model.
Exact Match Accuracy = (# instances with perfect match) / (total instances (n=250))
Label Accuracy = True Positive / (True Positive + False Positive + False Negative) = correct routes / all routes that appeared
Correctness Consistency = (instances with all 3 runs correct) / (total instances)

**Table 3.** Benchmarks 4-6: Drug-drug Interaction Identification, Renal Dose Identification, and Drug Indication Identification.

|  | **GPT-4o-mini (n=250)** | **MedGemma-27B (n=250)** | **LLaMA3-70B (n=250)** |
| --- | --- | --- | --- |
| ***Drug-drug Interaction Identification*** |  |  |  |
| Accuracy | 70.4% [64.8, 75.7] | 66.5% [61.2, 71.9] | 66.7% [61.1, 72.3] |
| Accuracy for Category C Interactions (n=81) | 62.1% [52.0, 72.4] | 56.8% [46.3, 67.1] | 57.6% [47.4, 68.0] |
| Accuracy for Category D Interactions (n=92) | 75.4% [66.3, 83.9] | 76.5% [67.1, 84.7] | 70.3% [60.7, 78.8] |
| Accuracy for Category X Interactions (n=77) | 73.2% [63.0, 82.2] | 64.9% [55.7, 75.2] | 71.9% [62.1, 81.9] |
| Correctness Consistency Score, n (%) | 169 (67.6) | 155 (62.0) | 160 (64.0) |
| ***Renal Dose Identification*** |  |  |  |
| F1 | 83.3% [77.6, 88.0] | 76.9% [70.8, 82.5] | 83.2% [77.5, 88.0] |
| Recall | 86.9% [80.3, 92.7] | 91.6% [86.3, 96.3] | 90.0% [84.1, 95.5] |
| Precision | 80.0% [72.5, 86.8] | 66.2% [58.5, 73.6] | 77.3% [69.7, 84.1] |
| Correctness Consistency Score, n (%) | 206 (82.4) | 186 (74.4) | 210 (84.0) |
| ***Drug Indication Identification*** |  |  |  |
| Accuracy | 96.5% [94.1, 98.5] | 96.9% [94.7, 98.9] | 97.6% [95.6, 99.2] |
| Correctness Consistency Score, n (%) | 244 (97.6) | 244 (97.6) | 247 (98.8) |

Values reported as mean [95% confidence interval] % of performance. Higher numbers indicate better performance
Accuracy = (True Positive + True Negative) / (True Positive + True Negative + False Positive + False Negative), this shows the fraction of correct predictions
Recall: true positive/ (true positive + false negative). Low recall indicates many false negatives.
Precision: true positive/ (true positive + false positive). Low precision indicates many false positives
F1: $2 \times (\text{precision} \times \text{recall}) / (\text{precision} + \text{recall})$ . Low F1 indicates many false negatives, false positives, or both.
Correctness Consistency = (instances with all 3 runs correct) / (total instances)
Drug interaction categories are defined as follows:
Category X — Avoid combination: This drug pair should generally not be used together because the clinical risk outweighs any potential benefit.
Category D — Consider therapy modification: The interaction is clinically relevant, and modification of therapy (dose adjustment, substitution, or precautions) should be considered.

Performance patterns diverged in safety-critical tasks. As shown in **Table 3**, GPT-4o-mini achieved the highest overall accuracy for drug–drug interaction classification (70.4% [95% CI: 64.8-75.7]) and the highest correctness-consistency score (67.6%). Although MedGemma-27B did not lead overall, it showed the strongest performance in Category D interactions (76.5% [95% CI: 67.1-84.7]), a clinically significant subclass that typically requires therapeutic modification.^14^ For renal dose adjustment identification, GPT-4o-mini and LLaMA3-70B had similar performance (F1 83.3% [95% CI: 77.6-88.0] and F1 82.3% [95% CI: 77.5-88.0], respectively).

Correctness-consistency scores varied widely across tasks, from 8.0% to 98.8%, reflecting differences in the stability of correct outputs across repeated prompt variants. These trends, detailed in **Table 2** and **Table 3**, show that consistency was highest in binary tasks such as drug-indication identification and lowest in multilabel tasks such as formulation and route identification. No model demonstrated uniformly superior consistency across all benchmarks.

A comparison of performance profiles across all six benchmarks is shown in **Figure 2**. LLaMA3-70B showed strong performance in structured medication-knowledge tasks requiring multilabel or compositional reasoning. GPT-4o-mini showed its greatest strength in safety-critical tasks involving drug–drug interactions and renal dose adjustments. MedGemma-27B performed competitively within selected interaction categories. These findings illustrate heterogeneous capability distributions among current LLMs and highlight the need for multi-domain evaluation when assessing medication-related clinical readiness.

**Figure 2.**
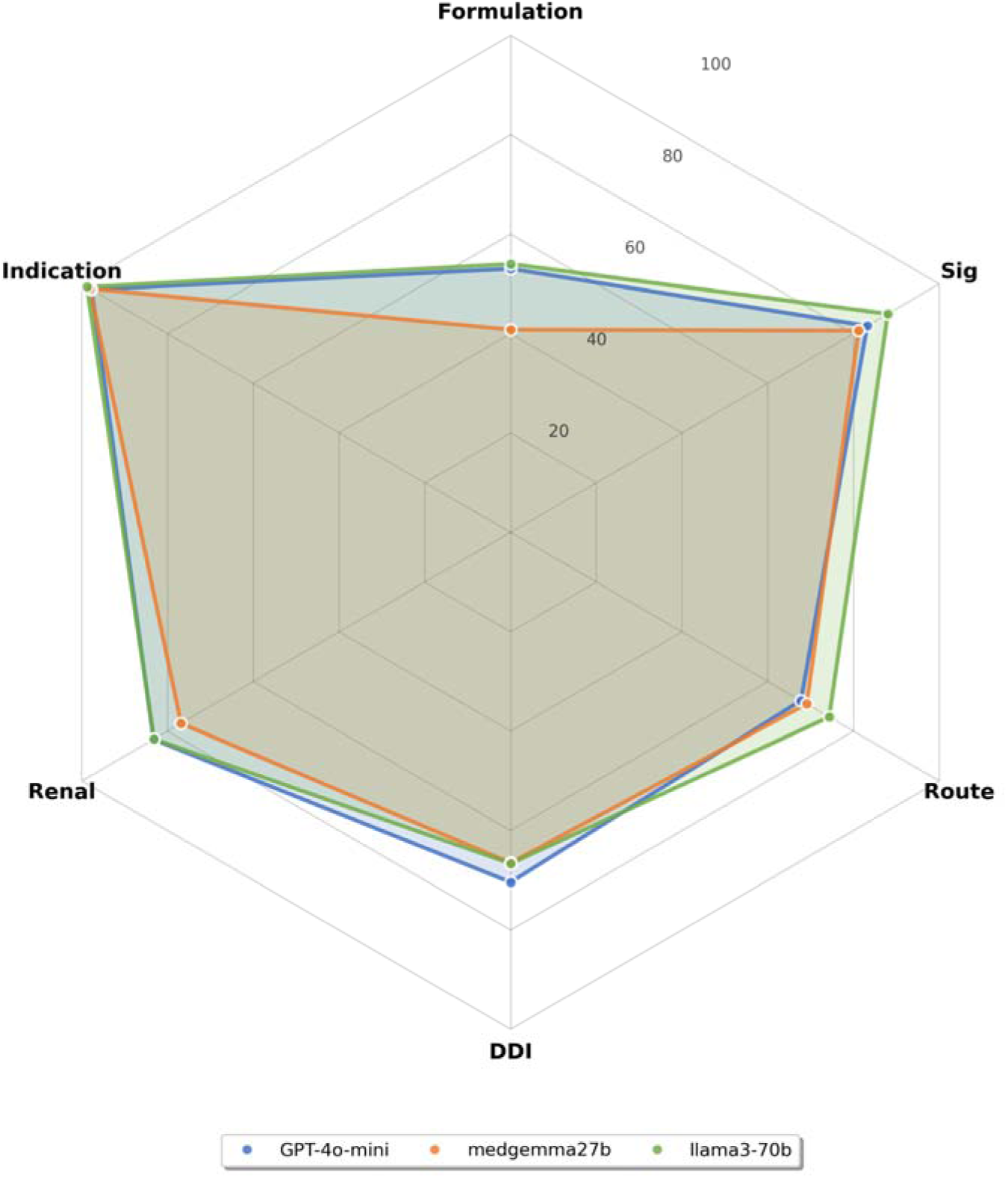
Comparison of 3 large language models across 6 medication benchmarks Accuracy is reported for indication, order generation, and drug-drug interaction benchmarks. F1 is reported for renal dose adjustment, route, and formulation benchmarks. Both metrics utilize the same scale, with 0 representing the worst value and 100 representing the best value. DDI: drug-drug interaction

## Discussion

We present six benchmark tasks as well as clinician-annotated datasets and standardized prompts that are available for use to test the safety and efficacy performance of LLMs for medication tasks. This evaluation marks the first time that a suite of benchmarking tasks has been specifically designed and annotated by clinicians for this use. This evaluation is an important step for the development of safe, transparent, and high-performing LLMs for pharmacy-related use because each benchmark task represents a discrete, real-world patient care task that must be performed with nearing perfect accuracy to avoid medication-related harm. While these tasks do not exhaustively represent the myriad of tasks related to the safe use of medications, this suite represents an important early indicator that an LLM could potentially have the appropriate knowledge, pre-training, and prompt engineering to be used for medication management.

One strength of this analysis was robust review of LLM outputs by clinicians and measurement of harm potential. The drug formulation benchmark yielded many inconsistencies, including a variety of false positives and false negatives. Less than 20% of drugs had all correct formulations provided in each run, although this information is readily available on several drug information references, including LexiDrug, and has capabilities to be programmed into EHRs.^14^ For the drug order generation benchmark, the prompt specifically limited order generation to oral routes in an attempt to decrease possibilities and complexity that may be seen with other routes. However, even with one of the simplest routes for order generation, error rates exceeded 10% for each model, with a range of harm scores. The drug route benchmark revealed significant inconsistencies, including high false positives. This was present for most drugs, including drugs with relatively few routes. The DDI benchmark yielded moderate performance from all models; however, several severe DDIs were missed. The renal dose adjustment benchmark yielded moderate performance as well, with interesting false positives and false negatives. The drug indication identification benchmark yielded the highest performance from all models; however, errors here would likely have resulted in patient harm as well. While the LLMs showed promise for completing medication-related benchmarking tasks, substantial performance improvements are needed as it relates to safety. Notable errors for the benchmarks include:

- Drug Order Generation: MedGemma-27B recommended in all runs to give dofetilide 12.5 mg by mouth twice daily. The correct dose is 125 mcg twice daily, so the recommended dose was a 100-fold overdose, which would have high potential to result in a life-threatening arrythmia (rated as a 4 on the HAMEC score).14,18 Additional errors that were classified as a 3 on the HAMEC score included all models recommending an underdose of flucytosine, a medication used to treat serious fungal infections, and an overdose of nitroglycerin from each model.14,18
- Drug Route Matching: amitriptyline is only available as oral but was listed as IV continuous, IV intermittent, intramuscular, oral, and topical by GPT-4o-mini; intramuscular, IV continuous, and oral by MedGemma-27B; and intramuscular and oral by LLaMA3-70B).14
- Drug-Drug Interaction: all models did not recognize the interaction between Andexanet alfa and heparin (a category X interaction) and carbapenems and valproic acid (a category D interaction that could result in seizures). Additionally, each model often reported only a partial drug name (such as “globulin” instead of “immune globulin” and “chloride” instead of “potassium chloride”), which could increase confusion.14 Interestingly, each model had 2-4 spelling errors where the medications within the DDIs were correct, but the LLM misspelled the medication, for example “mexelitine” instead of “mexiletine” and “cefpime” instead of “cefepime.”
- Renal Dose Adjustment: False positives included propranolol and rasburicase and false negatives included chlordiazepoxide and methylnaltrexone, which have clear dose adjustments in the package insert.14
- Drug Indication: GPT-4o-mini and MedGemma-27B both selected daptomycin as the medication associated with treatment of methicillin-resistant staphylococcus aureus (MRSA) pneumonia over ceftaroline; however, daptomycin is inactivated by pulmonary surfactant in the lungs and cannot be used for MRSA pneumonia.31 Additionally, LLaMA3-70B and MedGemma-27B selected valproic acid as the correct medication for the prevention of atrial fibrillation-related complications as opposed to argatroban, an anticoagulant. Giving valproic acid as opposed to an anticoagulant would have increased the risk of stroke.32 MedGemma-27B selected propranolol for thyrotoxicosis, which is correct, but propranolol was not on the answer list; metoprolol was the correct answer. This example of not following prompt instructions was also present in other benchmarks, notably the order generation benchmark where 1-7% of outputs did not follow prompt instructions.

Recent viewpoints have drawn parallels between education and AI benchmarking. Healthcare professions, including both medical and pharmacy schools, use a construct of competency-based medical education (CBME), in which entrustable professional activities or “EPAs” serve as observable and measurable units of practice that are teachable through modeling and feedback. These viewpoints suggest an AI-CBME approach that defines competencies and milestones in terms of EPAs.^19,20^ The six benchmarks are reflective of distinct tasks necessary for a broader EPA around the safe and responsible use of medication therapy.

The Three Principles is a previously proposed paradigm for safe medication use that include: (1) treatment of the underlying cause (e.g., an antibiotic for an infection), (2) provision of supportive care (e.g., an analgesic for pain secondary to the infection), and (3) avoidance of iatrogenic harm (e.g., avoidance of an antibiotic that the patient has a documented drug allergy for).^21,22^ While a seemingly simple paradigm that places the patient at the center of the process, medication harm remains a leading cause of morbidity and mortality.^23,24^ The high rates of medication errors, ADEs, polypharmacy, and other inappropriate prescribing are a motivation for the development of data-driven tools to make medication use safer; however, indiscriminate use of the general Internet (which contains high amounts of medication misinformation) and real-world electronic health record data (which certainly contains inappropriate prescriptions) has substantial risk for developing LLMs that provide unsafe medication recommendations. Indeed, rigorous clinician validation is absolutely essential for training and benchmarking datasets in this space. A recent example was DrugGPT,^25^ which included a DDI dataset without clinical validation that listed DDI pairs with directly conflicting / inaccurate information as well as outdated medications, including anticoagulants, which are one of the most dangerous and often misused medications on the market.^26^ An additional agent, MedAgentBench, creates a virtual electronic health record and can accomplish medication-related tasks such as ordering medications, but basic rules (for example, replacing potassium with a fixed linear dosing rule of 10mEq per 0.1mEq/L below 3.5) do not account for patient-specific factors (in this example, the patient’s renal function is not taken into account which could result in an excessive potassium replacement if using this linear dosing rule in a patient on dialysis) and may be marked as “correct” but have the potential to cause substantial patient harm.^27^ Recent studies have shown the fragility of LLMs in the face of poisoning (also known as adversarial attacks)^9,28^ and moreover even simple tasks like discriminating brand and generic drug names proved challenging.^29,30^

This study has several limitations: most notably, with just six tasks and 250 queries each, this series is not fully reflective of the complexities of the medication management process. Thus, an LLM could conceivably have high performance on these tasks and still show safety concerns in other domains. Moreover, given that prompt engineering and other methodologies can substantially improve performance, there is a possibility that with more advanced methodology, performance would have improved. Additionally, there are many general and specific-purpose LLMs that were not used in this study but are available and may have different performance than the sample LLMs included in this analysis. While these aspects were deemed outside of the scope of this foundational work, these are important future areas of exploration.

Ultimately, these LLMs failed to show consistent achievement of critically important medication management tasks. While LLMs have been able to pass licensing exams such as the Medical College Admission Test (MCAT) or United States Medical Licensing Examination (USMLE),^33,34^ they still are lacking the ability to consistently complete basic tasks like identify how medications can be given or what they are used for. This warrants tremendous concern about what basic functionality might be missing, regardless of how well an LLM can complete a standardized test.

## Conclusion

Six clinically validated medication task benchmark tasks were created to assess the five major categories (safety, adherence, operations, indication, effectiveness) of comprehensive medication management. The tested LLMs demonstrated inconsistent performance with accuracy or F1 scores ranging widely across benchmarks and LLMs. Future applications of these benchmarks could improve confidence in the safety of LLMs for clinical use.

## Data Availability

All data produced are available online at https://github.com/sikora07/AIChemist

## Acknowledgements

Christopher Barry

