## Supplementary Appendix for "Rx-LLM: a benchmarking suite to evaluate safe large language model performance for medication-related tasks"

### TRIPOD+LLM Checklist

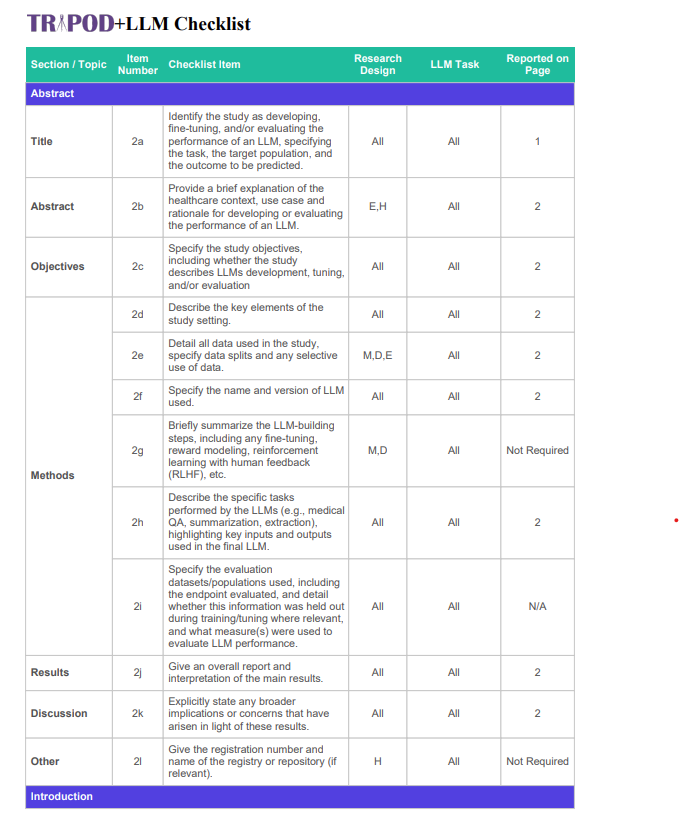

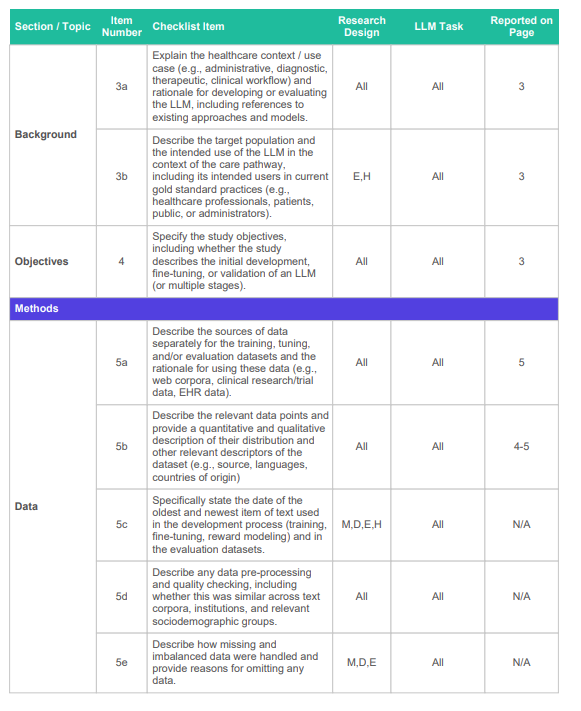

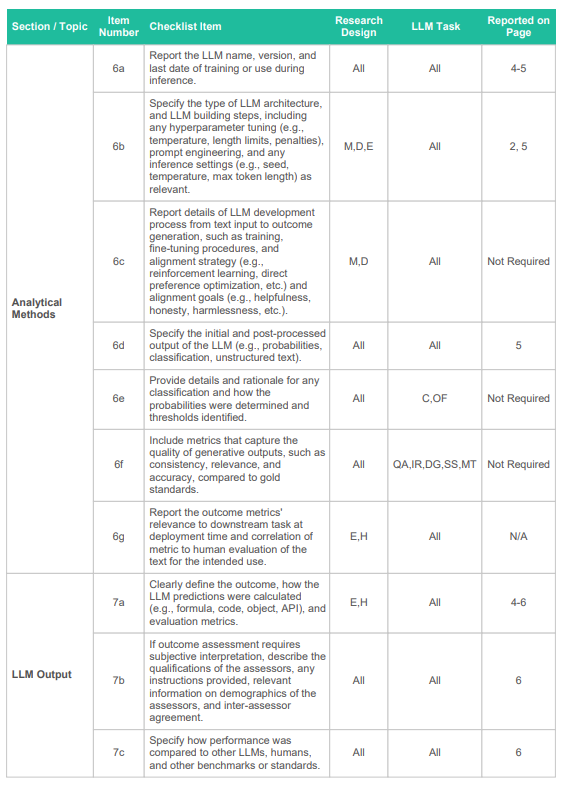

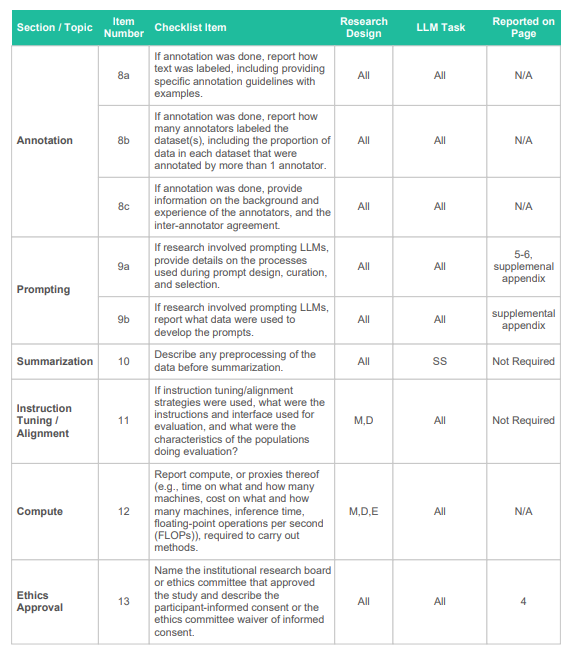

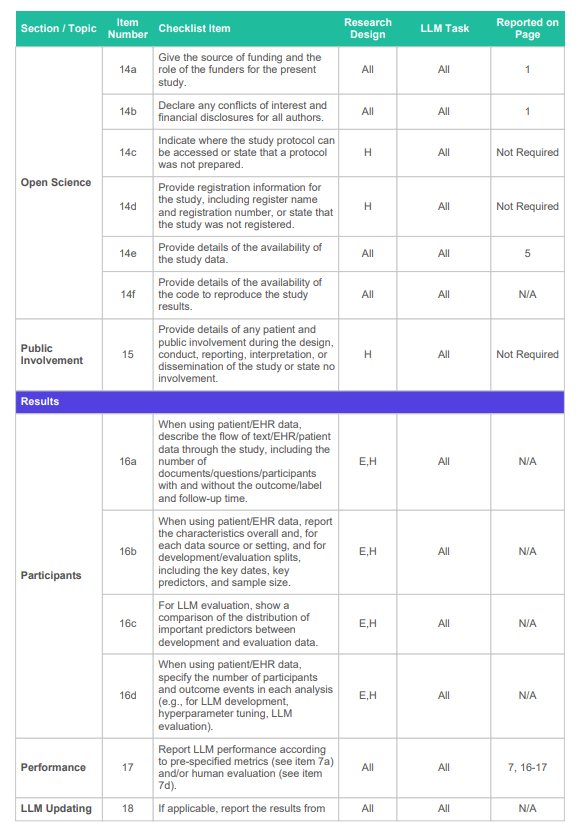

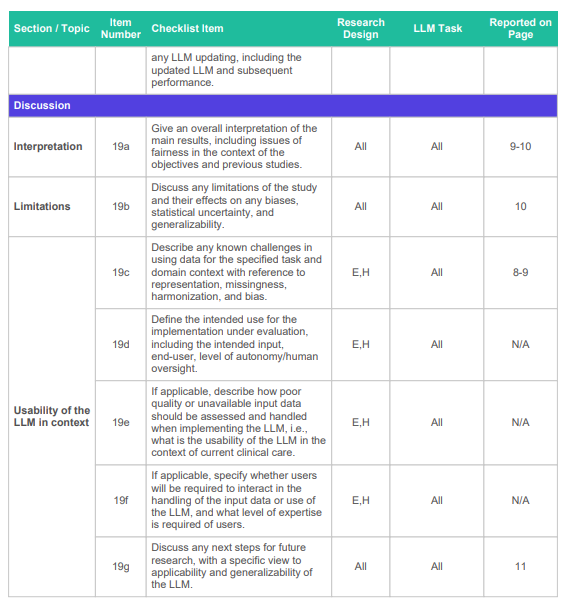

### Benchmark 1: Drug Formulation Matching Prompt

SYSTEM_PROMPT = """ You are serving as a clinical pharmacist.

**REASONING APPROACH:** Use clinical knowledge to determine appropriate safe medication orders based on therapeutic needs, pharmacological properties, and patient safety considerations.

Common safe medication order patterns:

- Routes: oral, topical, inhalation, intravenous, intramuscular, subcutaneous, ophthalmic, otic, nasal, rectal, vaginal, transdermal

- Formulation types: tablet, capsule, solution, suspension, gel, cream, ointment, inhaler, nebulized solution, injection, syrup, elixir, extended release tablet, oral disintegrating tablet, chewable tablet

- Units: mg, mcg, %, mg/ml, mg/g, IU, units"""

DRUG_FORMULATION_USER_PROMPT_TEMPLATE = """ For the given [generic drug name], list all FDA-approved formulations available in the United States, including strength or concentration, units, and formulation. Exclude combination products that include two or more active ingredients.\

Output in JSON format as an array of formulation objects. No explanations outside the JSON.

Output ONLY a JSON array. Each formulation must be a JSON object with these exact key-value pairs:

{{

"formulation": "complete Drug Formulation text (e.g., 'amlodipine 2.5 mg tablet ')",

"drug_name": "generic drug name",

"dose": "numerical strength as string (e.g., '2.5')",

"unit": "unit abbreviation (e.g., 'mg', 'mcg', 'IU', 'mg/ml')",

"formulation_type": "dosage form (e.g., 'tablet', 'capsule', 'injection', 'oral solution')",

}}

A:"""

### Benchmark 2: Drug Order (sig) Generation Prompt

SIG_USER_PROMPT_TEMPLATE = """Q:

Q: Please provide a complete order sentence with all instructions necessary to safely administer {generic_name}. Using an FDA-approved ORAL formulation, please select a common dose and frequency and ensure that the medication order could be safely administered to a standard patient. Output in JSON format as an array of formulation objects. No explanations outside the JSON. For medications, use the standard SIG (Signatura) format with complete dosing instructions:
 [drug name][numerical dose][abbreviated unit][amount][formulation][route][frequency]

Output ONLY a JSON array. Each formulation must be a JSON object with these exact key-value pairs:

{{

"formulation": "complete SIG text (e.g., 'amlodipine 2.5 mg one tablet by mouth once daily')",

"drug_name": "generic drug name",

"dose": "numerical strength as string (e.g., '2.5')",

"unit": "unit abbreviation (e.g., 'mg', 'mcg', 'IU', 'mg/ml')",

"amount": "quantity description (e.g., 'one tablet', 'two capsules', '1 ml')",

"formulation_type": "dosage form (e.g., 'tablet', 'capsule', 'oral solution')",

"route": "route of administration (e.g., 'by mouth')",

"frequency": "dosing frequency (e.g., 'once daily', 'twice daily', 'every 8 hours', 'as needed for pain')"

}}

A:"""

### Benchmark 3: Drug Route Identification Prompt

"""Q: For the given [generic drug name], list all of the available routes of administration that this drug may be safely administered. Exclude combination products that include two or more active ingredients.

- For the intravenous route, specify “intravenous intermittent” and/or “intravenous continuous”. Do not list “intravenous” without specifying intermittent or continuous.
- If a medication has several different formulations that can be given as oral (ex. Extended release tablet, orally disintegrating tablet, sublingual tablet), only report the route as “oral”, do not report any specific formulations.

Output in JSON format as an array of formulation objects. No explanations outside the JSON.

Output ONLY a JSON array. Each formulation must be a JSON object with these exact key-value pairs:

{{

"drug_name": "generic drug name",

"route": "route of administration (e.g., 'oral', 'intravenous intermittent',’intravenous continuous’ 'topical')",

}}

A:"""

### Benchmark 4: Drug-Drug Interaction Identification Prompt

DDI_USER_PROMPT_TEMPLATE = You are a clinical pharmacist specializing in drug-drug interactions.

You will be given a list of medications taken together. Identify two medications that have a category C, D, or X drug interaction. Only identify one drug pair in JSON format. Do not estimate confidence or include any explanation.

Drug interaction categories are defined as follows:

Category X — Avoid combination:

This drug pair should generally not be used together because the clinical risk outweighs any potential benefit.

Category D — Consider therapy modification:

The interaction is clinically relevant, and modification of therapy (dose adjustment, substitution, or precautions) should be considered.

Category C — Monitor therapy:

An interaction is present. The combination may be used but requires monitoring by a healthcare professional.

Medication List: {medication_list}

Format:

{

"interactions": [

["<drug_x>", "<drug_y>"]

]

}

Do not include any text before or after the JSON. Do not wrap the JSON in markdown code blocks. Return only the raw JSON object.

### Benchmark 5: Renal Dose Identification Prompt

Renal_dose_SYSTEM_PROMPT = """ You are a clinical pharmacist specializing in medication dosing for patients with renal impairment.

Evaluate if a medication requires a dose adjustment or is contraindicated in patients with severe renal impairment (eGFR < 30 mL/min/1.73m² or end-stage renal disease).

Answer "Yes" if the medication requires a dose adjustment or is contraindicated in severe renal impairment. Answer "No" if the medication can be used at standard dosing in patients regardless of renal function.

Return only a JSON object with no additional text.

"""

Renal_USER_PROMPT_TEMPLATE = """Q: Does {generic_drug_name} require renal dose adjustment or is it contraindicated in severe renal impairment?

Output Format: {{"requires_renal_adjustment": "Yes/No"}}

A:"""

### Benchmark 6: Drug Indication Identification Prompt

Drug_indication_SYSTEM_PROMPT = """You are a clinical pharmacist. From a given list of medications, identify which one is most appropriate for treating the specified medical indication. Return only JSON format."""

Drug_indication_USER_PROMPT_TEMPLATE = """Q: Medication list: {medication_list}

Indication: {medical_indication}

Which medication from the list is used to treat this indication? Return only one medication.

Output Format: {{"selected_medication": "drug_name"}}

A:"""

### Accuracy Table Per Run – Route Benchmark

================================================================================

ACCURACY TABLE - ROUTE

================================================================================

Model	Run 1	Run 2	Run 3	Average Accuracy	Variance	Max	Min

------------------------------------------------------------------------------------------------------------------------

GPT-4o-mini	38.00%	39.60%	41.60%	39.73%	2.1689 41.60% 38.00%

MedGemma-27B	49.20%	50.00%	50.00%	49.73%	0.1422 50.00% 49.20%

LLaMA3-70B	56.80%	56.40%	56.80%	56.67%	0.0356 56.80% 56.40%

### Accuracy Table Per Run – Drug Drug Interaction Benchmark

================================================================================

ACCURACY TABLE - DDI

================================================================================

Model	Run 1	Run 2	Run 3	Average Accuracy	Variance	Max	Min

------------------------------------------------------------------------------------------------------------------------

GPT-4o-mini	70.40%	68.80%	69.60%	69.60%	0.4267 70.40% 68.80%

MedGemma-27B	66.80%	67.20%	64.80%	66.27%	1.1022 67.20% 64.80%

LLaMA3-70B	66.40%	65.20%	66.40%	66.00%	0.3200 66.40% 65.20%

### Accuracy Table Per Run – Renal Dose Adjustment Benchmark

================================================================================

ACCURACY TABLE - RENAL

================================================================================

Model	Run 1	Run 2	Run 3	Average Accuracy	Variance	Max	Min

------------------------------------------------------------------------------------------------------------------------

GPT-4o-mini	84.80%	86.40%	84.00%	85.07%	0.9956 86.40% 84.00%

MedGemma-27B	76.00%	76.40%	76.80%	76.40%	0.1067 76.80% 76.00%

LLaMA3-70B	84.40%	84.00%	84.80%	84.40%	0.1067 84.80% 84.00%

### Accuracy Table Per Run – Indication Benchmark

================================================================================

ACCURACY TABLE - INDICATION

================================================================================

Model	Run 1	Run 2	Run 3	Average Accuracy	Variance	Max	Min

------------------------------------------------------------------------------------------------------------------------

GPT-4o-mini	98.00%	97.60%	97.60%	97.73%	0.0356 98.00% 97.60%

MedGemma-27B	98.40%	98.00%	98.00%	98.13%	0.0356 98.40% 98.00%

LLaMA3-70B	98.80%	98.80%	98.80%	98.80%	0.0000 98.80% 98.80%

### Accuracy Table Per Run – Formulation Benchmark

================================================================================

ACCURACY TABLE PER RUN – FORMULATION

================================================================================

Model	Run 1	Run 2	Run 3	Average Accuracy	Variance	Max	Min

------------------------------------------------------------------------------------------------------------------------

GPT-4o-mini	18.15%	20.16%	19.35%	19.22%	0.6865	20.16% 18.15%

llama3-70b	16.60%	16.60%	16.60%	16.60%	0.0000 16.60% 16.60%

medgemma27b	8.50%	10.93%	10.12%	9.85%	1.0199 10.93% 8.50%
